# Machine-Learning Model Identifies New Diagnostic Criteria for Beckwith-Wiedemann Spectrum

**DOI:** 10.64898/2026.06.22.26355886

**Authors:** Sylvie A. Adams, Aravind Viswanathan, Bamelak T. Duki, Andrew M. George, Jill A. Fahrner, Darko Stefanovski, Christopher M. Cielo, Jennifer M. Kalish

**Author notes:** **Address correspondence to:** Jennifer M. Kalish, MD, PhD, Division of Human Genetics, Children’s Hospital of Philadelphia, 3501 Civic Center Blvd, CTRB 3028, Philadelphia PA, 19104 USA.

## Abstract

**Objective:** Beckwith-Wiedemann spectrum (BWSp) is an overgrowth and cancer predisposition disorder caused by genetic and epigenetic alterations of chromosome 11p15. The 2018 international consensus produced a clinical scoring system to capture the phenotypic variability of BWSp and guide genetic testing and clinical management, including tumor screening, in patients without molecular confirmation. In this study, we evaluated BWSp predictors to identify the most informative features.

**Methods:** Supervised machine learning analyzed 25 phenotypic features in 555 patients with BWSp and 150 controls. Logistic regression, combined with a purposeful stepwise selection algorithm, identified a subset of features that can accurately classify subjects. Model performance was evaluated in a testing set and validated externally.

**Results:** The final model included six predictors: macroglossia, lateralized overgrowth, midface flattening, hepatomegaly, omphalocele, and developmental delay. Developmental delay was the only negative predictor; macroglossia (OR 46.10) and lateralized overgrowth (OR 27.87) were the strongest predictors. The proposed model and 2018 system did not differ in classification performance for testing (*P* = .39) or external (*P* = .15) sets.

**Conclusion:** A simplified diagnostic model, driven by macroglossia and lateralized overgrowth, differentiates between patients with BWSp and controls with performance comparable to the 2018 system.

And may help physicians prioritize BWSp evaluation.

## Introduction

Beckwith-Wiedemann spectrum (BWSp) (OMIM 130650) is the most common epigenetic overgrowth and cancer predisposition condition, affecting nearly 1 in 10,000 patients.^1^ BWSp is caused by epigenetic and genetic alterations of chromosome 11p15. Common molecular mechanisms include loss of methylation at imprinting cluster 2 (IC2 LOM), paternal uniparental isodisomy of chromosome 11 (pUPD11), gain of methylation at imprinting cluster 1 (IC1 GOM), and pathogenic variants in *CDKN1C*. In addition, structural alterations involving 11p15 account for about 5% of cases.^2^ Patients with IC2 LOM, pUPD11, IC1 GOM, and 11p15 anomalies have increased risks of hepatoblastoma and Wilms tumor; patients with pathogenic *CDKN1C* variants have an increased risk of neuroblastoma.^3,4^

The 2018 international consensus proposed a weighted clinical scoring system to capture the phenotypic variability of BWSp.^2^ This system includes cardinal (e.g., macroglossia, lateralized overgrowth, hyperinsulinism) and suggestive (e.g., ear creases/pits, facial nevus simplex, organomegaly) features, assigning two points per cardinal feature and one point per suggestive feature. Scores ≥ 4 and/or lateralized overgrowth support a clinical BWSp diagnosis in the absence of molecular confirmation.^2,5^

Phenotypic heterogeneity, mosaicism, and limited access to sensitive genetic testing complicate diagnosis of BWSp. Diagnostic guidelines play a key role in determining when to pursue genetic testing and if tumor screening is warranted without molecular confirmation. Machine learning provides an opportunity to evaluate the current guidelines, which were primarily informed by frequencies of features in the patient population and clinical expertise, and develop a simplified predictive model. Here, we present a statistically rigorous assessment of BWSp predictors to identify the most clinically informative features.

## Methods

### Data collection

Retrospective data abstraction was performed for a cohort of 555 patients with molecularly confirmed BWSp and 150 controls, all of whom were evaluated or had records reviewed at the Children’s Hospital of Philadelphia (CHOP) with the exception of 27 BWSp cases reported in the literature.^6,7^ Controls were patients dismissed from the overgrowth genetics clinic at CHOP after receiving evaluation for BWSp or another genetic syndrome.

BWSp diagnosis was confirmed with molecular testing performed on various tissues, including blood, skin, and tongue. Patients with positive results only in tumor and/or affected pancreas, but negative results in adjacent normal tissue, were excluded. Molecular subtypes were classified as IC2 LOM, IC1 GOM, pUPD11, genome-wide paternal uniparental isodisomy (GWpUPD), *CDKN1C*, and 11p15 anomalies. Methylation and mosaicism analyses were performed according to standard methodologies.^8^ BWSp features were assessed using established phenotyping criteria, and clinical scores were calculated using guidelines from the 2018 international consensus.^2,9^ A clinical BWSp diagnosis was defined as (1) a score ≥ 4 with at least one cardinal feature present and/or (2) lateralized overgrowth. Informed consent, where applicable, was obtained from participants or families according to the relevant IRB protocol. Chart review and data abstraction were performed under IRB13-010658 and IRB19-016459.

A collaborator (JAF) at Johns Hopkins University School of Medicine provided an independent, de-identified cohort for external validation of the final model. This collaborator did not have access to the model during development, testing, or validation. The external cohort comprised 10 patients with BWSp, 10 controls with a non-overgrowth genetic diagnosis, and 10 patients with a non-BWSp overgrowth condition.

### Statistical analysis

Descriptive and complementary analyses were performed in R v4.6.0. Categorical variables are summarized as counts (%) and continuous variables as mean (SD). Two pre-specified pairwise comparisons (IC1 GOM vs IC2 LOM; IC1 GOM vs pUPD11) were conducted to evaluate associations between (1) molecular subtype and blood-positive testing for BWSp, and (2) molecular subtype and meeting the 2018 clinical diagnostic criteria for BWSp (Yes/No). For each 2x2 comparison, Pearson’s chi-square test or Fisher’s exact test was used, as appropriate. These comparisons were pre-specified to assess whether the IC1 GOM subtype has reduced diagnostic yield on blood-based testing and/or reduced likelihood of meeting the 2018 diagnostic criteria, consistent with prior reports.^9–11^ P values were adjusted for multiple comparisons using the Holm method.

Model development was performed in Stata v18.0. Patients were randomly split into development (70%) and testing (30%) sets, stratified by BWSp status. Differences in categorical variables between these sets were assessed with chi-square or Fisher’s exact tests, as appropriate. Gestational age was compared using Welch’s t-test to account for unequal variances and unequal group sizes. Likewise, comparisons between development and external cohorts were performed with chi-square or Fisher’s exact tests, as appropriate, to describe cohort similarity. Statistical significance was defined as *P* < .05.

The logistic regression classifier was developed in four steps. First, Firth univariate logistic regression was used to establish which features showed a trend of statistical association (*P* < .50) with the outcome of interest (BWSp status) defined as a binary variable (Yes/No). Firth was used to address the possibility that some independent variables might provide perfect prediction of BWSp, which cannot be estimated with univariate logistic regression. The statistician was blinded to the features, other than that they were either continuous or categorical variables. Second, the independent variables identified in the previous step were subjected to a purposeful stepwise backward and forward algorithm to select a subset of features that were significantly associated (*P* < .05) with the outcome of interest. Third, the final multivariable model was estimated, and optimization of the cutoff probability was conducted for the estimation of sensitivity and specificity. Fourth and final, features (independent variables) that were rarely obtained were removed from the model. Internal validation to assess the stability of the model was conducted with bootstrapping. Using nonparametric receiver operating characteristic (ROC) analysis, model performance was evaluated in the testing set and subsequently validated in the external set, with results compared to the 2018 diagnostic system.

## Results

### Cohort description

The BWSp cohort included 555 molecularly confirmed patients: 315 females, 234 males, and 6 cases from the literature with sex not specified.^7^ Most patients resided in the United States. Common molecular subtypes were IC2 LOM (53.2%), pUPD11 (25.4%), and IC1 GOM (12.1%); less common subtypes included 11p15 anomalies (4.9%), *CDKN1C* variants (2.5%), and GWpUPD (1.6%) (Supplemental Table S1). The most frequent clinical features were macroglossia, lateralized overgrowth, ear creases/pits, and facial nevus simplex (Table 1).

**Table 1.** Clinical Features of the BWSp Cohort^a^.

|  | Total<sup>b</sup> | IC1 GOM | IC2 LOM | pUPD11 | CDKN1C | GWpUPD | 11p15 anomaly |
| --- | --- | --- | --- | --- | --- | --- | --- |
| **Cardinal Features** |  |  |  |  |  |  |  |
| Macroglossia<sup>c</sup> | 428/555 (77.1) | 29/67 (43) | 268/295 (90.8) | 87/141 (62) | 14/14 (100) | 1/9 (11) | 27/27 (100) |
| Omphalocele<sup>c</sup> | 92/555 (16.6) | 0/67 (0) | 73/295 (24.7) | 10/141 (7) | 5/14 (36) | 1/9 (11) | 3/27 (11) |
| Lateralized overgrowth<sup>c</sup> | 383/555 (69.0) | 50/67 (75) | 182/295 (61.7) | 131/141 (93) | 3/14 (21) | 5/9 (56) | 11/27 (41) |
| Multifocal/bilateral WT/NB<sup>c</sup> | 14/555 (2.5) | 9/67 (13) | 0/295 (0) | 4/141 (3) | 0/14 (0) | 0/9 (0) | 1/27 (4) |
| Hyperinsulinism<sup>c</sup> | 96/555 (17.3) | 8/67 (12) | 28/295 (9.5) | 43/141 (30) | 2/14 (14) | 7/9 (78) | 8/27 (30) |
| Pathology findings<sup>c,d</sup> | 17/505 (3.4) | 0/61 (0) | 4/275 (1.5) | 7/117 (6) | 2/14 (14) | 2/9 (22) | 2/27 (7) |
| **Suggestive Features** |  |  |  |  |  |  |  |
| Large for gestational age<sup>c</sup> | 207/551 (37.6) | 25/67 (37) | 106/295 (35.9) | 53/137 (39) | 3/14 (21) | 1/9 (11) | 18/27 (67) |
| Facial nevus simplex<sup>c</sup> | 264/528 (50.0) | 11/64 (17) | 195/283 (68.9) | 38/129 (29) | 8/14 (57) | 1/9 (11) | 9/27 (33) |
| Polyhydramnios<sup>c</sup> | 137/543 (25.2) | 11/65 (17) | 79/289 (27.3) | 26/137 (19) | 4/14 (29) | 3/9 (33) | 14/27 (52) |
| Placentomegaly | 50/522 (9.6) | 2/61 (3) | 31/281 (11.0) | 12/129 (9) | 1/13 (8) | 2/9 (22) | 2/27 (7) |
| Ear creases/pits<sup>c</sup> | 325/555 (58.6) | 22/67 (33) | 199/295 (67.5) | 68/141 (48) | 11/14 (79) | 5/9 (56) | 19/27 (70) |
| Transient hypoglycemia<sup>c</sup> | 190/554 (34.3) | 16/67 (24) | 121/294 (41.2) | 39/141 (28) | 2/14 (14) | 1/9 (11) | 9/27 (33) |
| Tumor<sup>e</sup> | 65/555 (11.7) | 29/67 (43) | 9/295 (3.1) | 22/141 (16) | 0/14 (0) | 1/9 (11) | 4/27 (15) |
| Wilms tumor<sup>f</sup> | 40/555 (7.2) | 27/67 (40) | 2/295 (0.7) | 7/141 (5) | 0/14 (0) | 0/9 (0) | 4/27 (15) |
| Hepatoblastoma<sup>c</sup> | 21/555 (3.8) | 2/67 (3) | 6/295 (2.0) | 12/141 (9) | 0/14 (0) | 1/9 (11) | 0/27 (0) |
| Organomegaly |  |  |  |  |  |  |  |
| Nephromegaly<sup>c</sup> | 91/548 (16.6) | 15/67 (22) | 29/290 (10.0) | 31/139 (22) | 2/14 (14) | 2/9 (22) | 12/27 (44) |
| Hepatomegaly<sup>c</sup> | 102/548 (18.6) | 8/67 (12) | 45/290 (15.5) | 32/139 (23) | 2/14 (14) | 8/9 (89) | 7/27 (26) |
| Abdominal wall defect |  |  |  |  |  |  |  |
| Umbilical hernia<sup>c</sup> | 203/554 (36.6) | 15/67 (22) | 109/294 (37.1) | 63/141 (45) | 2/14 (14) | 2/9 (22) | 11/27 (41) |
| Diastasis recti<sup>c</sup> | 134/553 (24.2) | 15/67 (22) | 70/294 (23.8) | 34/141 (24) | 2/13 (15) | 2/9 (22) | 10/27 (37) |
| **Other Features** |  |  |  |  |  |  |  |
| Assisted reproductive technology<sup>c</sup> | 104/449 (23.2) | 10/52 (19) | 78/246 (31.7) | 11/114 (10) | 0/10 (0) | 0/5 (0) | 3/20 (15) |
| Midface flattening<sup>c</sup> | 159/505 (31.5) | 12/60 (20) | 108/270 (40.0) | 27/127 (21) | 5/11 (45) | 1/9 (11) | 5/26 (19) |
| Thin upper lip<sup>c</sup> | 146/505 (28.9) | 11/60 (18) | 95/270 (35.2) | 29/127 (23) | 4/11 (36) | 1/9 (11) | 6/26 (23) |
| Cardiac anomalies<sup>c</sup> | 166/549 (30.2) | 8/67 (12) | 91/291 (31.3) | 40/139 (29) | 4/14 (29) | 6/9 (67) | 16/27 (59) |
| Developmental delay<sup>c</sup> | 248/524 (47.3) | 20/62 (32) | 149/282 (52.8) | 53/129 (41) | 8/14 (57) | 3/8 (38) | 13/27 (48) |
| Prematurity (< 37 weeks)<sup>c</sup> | 219/530 (41.3) | 17/63 (27) | 130/282 (46.1) | 38/133 (29) | 8/14 (57) | 9/9 (100) | 17/27 (63) |
| Gestational age, mean (SD), weeks<sup>c</sup> | 36.4 (3.7) | 37.7 (3.3) | 36.0 (3.7) | 37.6 (2.9) | 34.5 (4.6) | 30.5 (3.7) | 35.0 (3.9) |

Abbreviations: WT/NB, Wilms tumor or nephroblastomatosis.
<sup>a</sup> Categorical variables reported as n/N (%), where N excludes patients with unknown or unavailable data.
<sup>b</sup> The total cohort of 555 patients includes two “other” molecular subtypes: one patient with IC2 LOM/IC1 LOM (BWSp/Russell-Silver syndrome) and one patient with IC1 GOM/CDKN1C. The data from these two patients are included in the total column but are not summarized in their own subtype column because the feature counts are low.
<sup>c</sup> Feature proceeded to stepwise selection step of model development.
<sup>d</sup> Placental mesenchymal dysplasia, adenomatosis of the pancreas, or cytomegaly of the adrenal glands.
<sup>e</sup> Tumors observed in this cohort include hepatoblastoma, neuroblastoma, multifocal/bilateral Wilms tumor, and unilateral Wilms tumor.
<sup>f</sup> Wilms tumor category includes both multifocal/bilateral and unilateral cases.

Patients with IC2 LOM (*P* < .001) and pUPD11 (*P* = .03) were more likely to test positive in blood than patients with IC1 GOM. Compared with the IC1 GOM group, IC2 LOM (*P* = .04) and pUPD11 (*P <* .001) subtypes were more likely to have enough clinical features for a clear clinical diagnosis of BWSp.

The control cohort (n = 150) comprised 71 females and 79 males; all controls resided in the United States. This cohort included 128 patients without a genetic diagnosis, 13 patients with a non-overgrowth genetic concern, and 9 patients with a non-BWSp overgrowth condition. Phenotypic characteristics for controls are summarized in Supplemental Table S2.

### Model development and performance

Model development, testing, and validation are delineated in Figure 1. After random stratification by BWSp status, development (389 BWSp, 105 controls) and testing (166 BWSp, 45 controls) sets had identical class distributions (79% BWSp, 21% controls). Comparisons between development and testing sets for BWSp cases and controls are shown in Supplemental Tables S3 and S4, respectively. For patients with BWSp, there were no significant differences between development and testing sets. Among controls, the development and testing sets diverged with respect to ear creases/pits, diastasis recti, and gestational age (*P* < .05), though mean gestational age only differed by 4.9 days and the frequency of prematurity was not significantly different.

**Figure 1.**
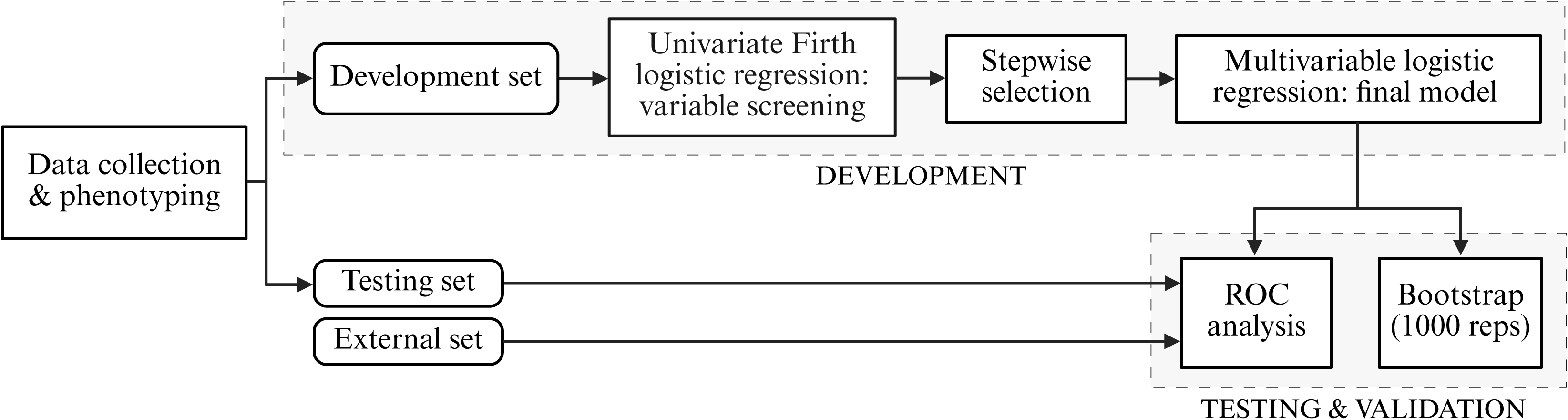
Flow diagram of model development, testing, and validation. Univariate logistic regression and stepwise selection identified a subset of predictors to undergo multivariable logistic regression and produce a final model. Receiver operating characteristic (ROC) analysis was used for model testing and validation; bootstrapping assessed model stability. Created with BioRender.com

Using univariate logistic regression, several features were omitted before moving forward with stepwise selection because their coefficients were not estimable from the data (Table 1). Stepwise selection identified a smaller subset of features that were significantly associated (*P* < .05) with BWSp status: nephromegaly, macroglossia, midface flattening, lateralized overgrowth, ear creases/pits, pathology findings, developmental delay, and facial nevus. Although stepwise selection initially removed hepatomegaly and omphalocele, they were reintroduced to the model due to their strong association with BWSp (OR∼>3, *P* < .01).

The final logistic model used six predictors: macroglossia, lateralized overgrowth, midface flattening, hepatomegaly, omphalocele, and developmental delay (Table 2). Developmental delay was the only negative predictor (presence of the factor indicates reduced likelihood of BWSp). Macroglossia and lateralized overgrowth were the strongest predictors, with macroglossia having a higher odds ratio (OR 46.10 [95% CI, 18.64-114.02], *P* < .001) than lateralized overgrowth (OR 27.87 [95% CI, 12.16-63.85], *P* < .001). Internal validation demonstrated the model’s stability: across 1000 bootstrap replicates the model failed to converge 37 times.

**Table 2.** Multivariable Odds Ratios for Features in the Proposed Model.

| Feature | OR (95% CI) | P value |
| --- | --- | --- |
| Macroglossia | 46.10 (18.64-114.02) | < .001 |
| Lateralized overgrowth | 27.87 (12.16-63.85) | < .001 |
| Midface flattening | 5.76 (1.37-24.31) | .02 |
| Hepatomegaly | 3.77 (1.11-12.80) | .03 |
| Omphalocele | 3.08 (0.69-13.71) | .14 |
| Developmental delay | 0.39 (0.18-0.83) | .01 |

In the testing set, the proposed model’s area under the curve (AUC) in ROC analysis was 0.90 (95% CI, 0.86-0.95), compared with 0.88 (95% CI, 0.83-0.94) for the 2018 diagnostic scoring system (Figure 2). AUC comparison showed that the proposed model and the 2018 diagnostic system did not significantly differ with respect to their classification performance (*P* = .39). To estimate the model’s sensitivity and specificity, we used a cutoff probability of 0.80. At this cutoff, the model correctly classified 90% of individuals in the testing set (sensitivity 90%, specificity 91%).

**Figure 2.**
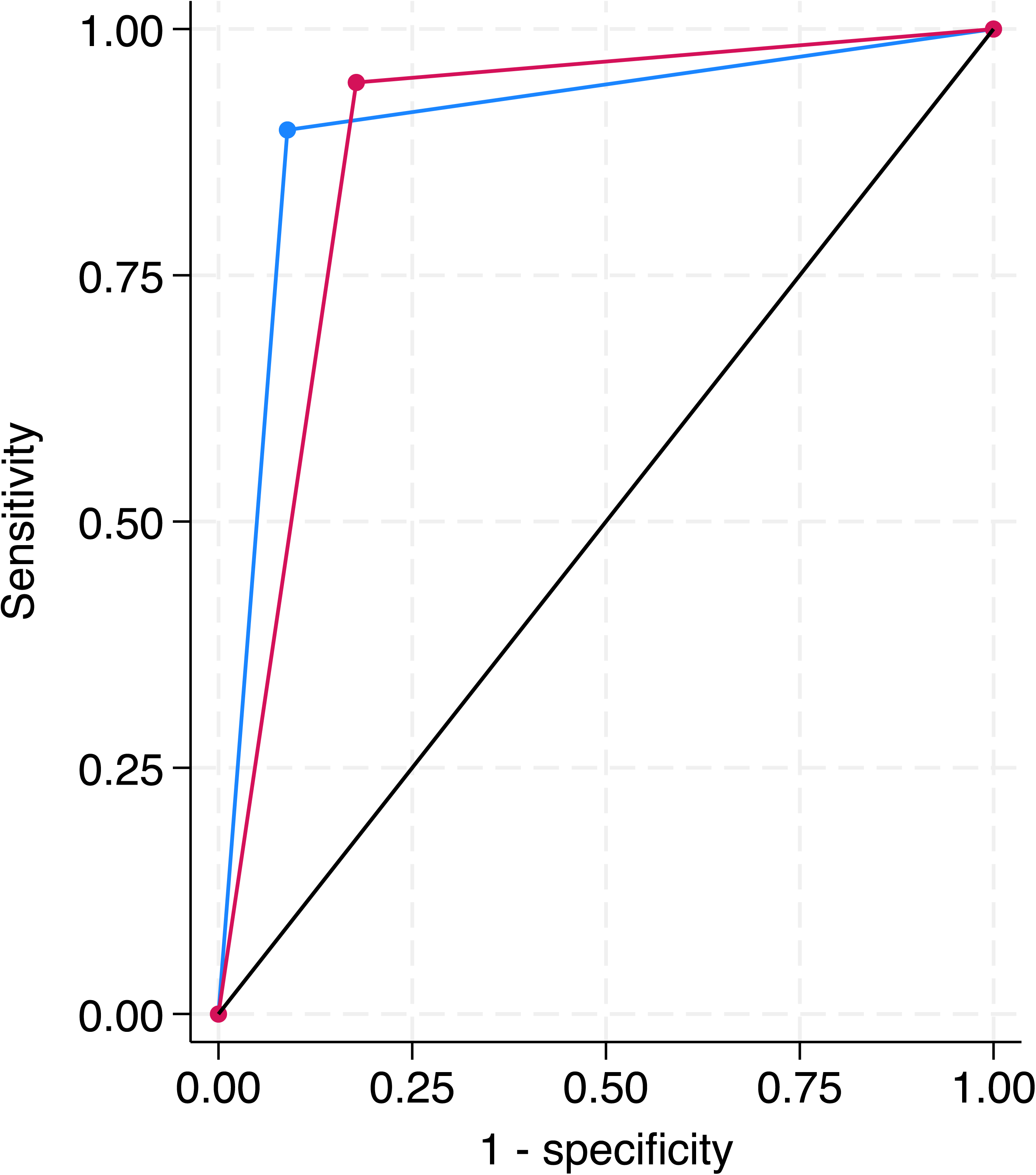
Receiver operating characteristic analysis with area under the curve (AUC) for proposed model and 2018 system: testing set (166 BWSp cases, 45 controls). Proposed model AUC = 0.90 (95% CI, 0.86-0.95, blue line); 2018 system AUC = 0.88 (95% CI, 0.83-0.94, red line). Classification performance was not significantly different (χ² = 0.73, *P* = .39).

The proposed six-feature model misclassified 17 of 166 BWSp cases in the testing set (Supplemental Table S5). Three patients were tested for BWSp due to presentation with a tumor, two of whom had both the IC1 GOM subtype and bilateral Wilms tumors. Of the eight subjects for whom suspicion of BWSp was prompted by presentation with a cardinal feature (other than tumors), two were initially evaluated for macroglossia, three for lateralized overgrowth, one for omphalocele, and two for hyperinsulinism. All missed subjects who had macroglossia or lateralized overgrowth also had developmental delay, which decreased the calculated probability of the individual having BWSp under the proposed model. The two patients whose original assessment is attributed to presenting with hyperinsulinism both had pUPD11. The missed patient with GWpUPD had a complex medical history from birth and underwent testing partly due to suspicion of a tumor, though ultimately no tumor was found. Other indications for testing in the remaining false negative cases are described in Supplemental Table S5.

### Model validation

The external validation cohort included 10 BWSp cases (5 IC2 LOM, 5 pUPD11), 10 controls with a non-overgrowth genetic diagnosis, and 10 patients with a non-BWSp overgrowth condition. Phenotypic characteristics of this cohort are summarized in Supplemental Table S6. Comparisons between development and external sets for BWSp cases and controls are shown in Supplemental Tables S7 and S8, respectively. For patients with BWSp, the external set had significantly higher frequencies of omphalocele, pathology findings, polyhydramnios, midface flattening, and cardiac anomalies (*P* < .05), which may reflect the small size of the external cohort and its restriction to IC2 LOM and pUPD11 subtypes. External controls had significantly higher frequencies of transient hypoglycemia, midface flattening, thin upper lip, and developmental delay when compared to development controls (*P* < .05). These between-cohort differences indicate that the external and development sets are not entirely similar, potentially accounting for differences in model performance.

The model correctly classified all cases in the external set. By comparison, the 2018 system successfully predicted BWSp status for all BWSp cases and controls but misclassified two patients with another overgrowth condition. In the external set, AUC was 1.00 (95% CI, 1.00-1.00) for the proposed model and 0.95 (95% CI, 0.88-1.00) for the 2018 diagnostic system (Figure 3). AUC comparison showed that these diagnostic systems did not significantly differ with respect to their classification performance in this data set (*P* = .15).

**Figure 3.**
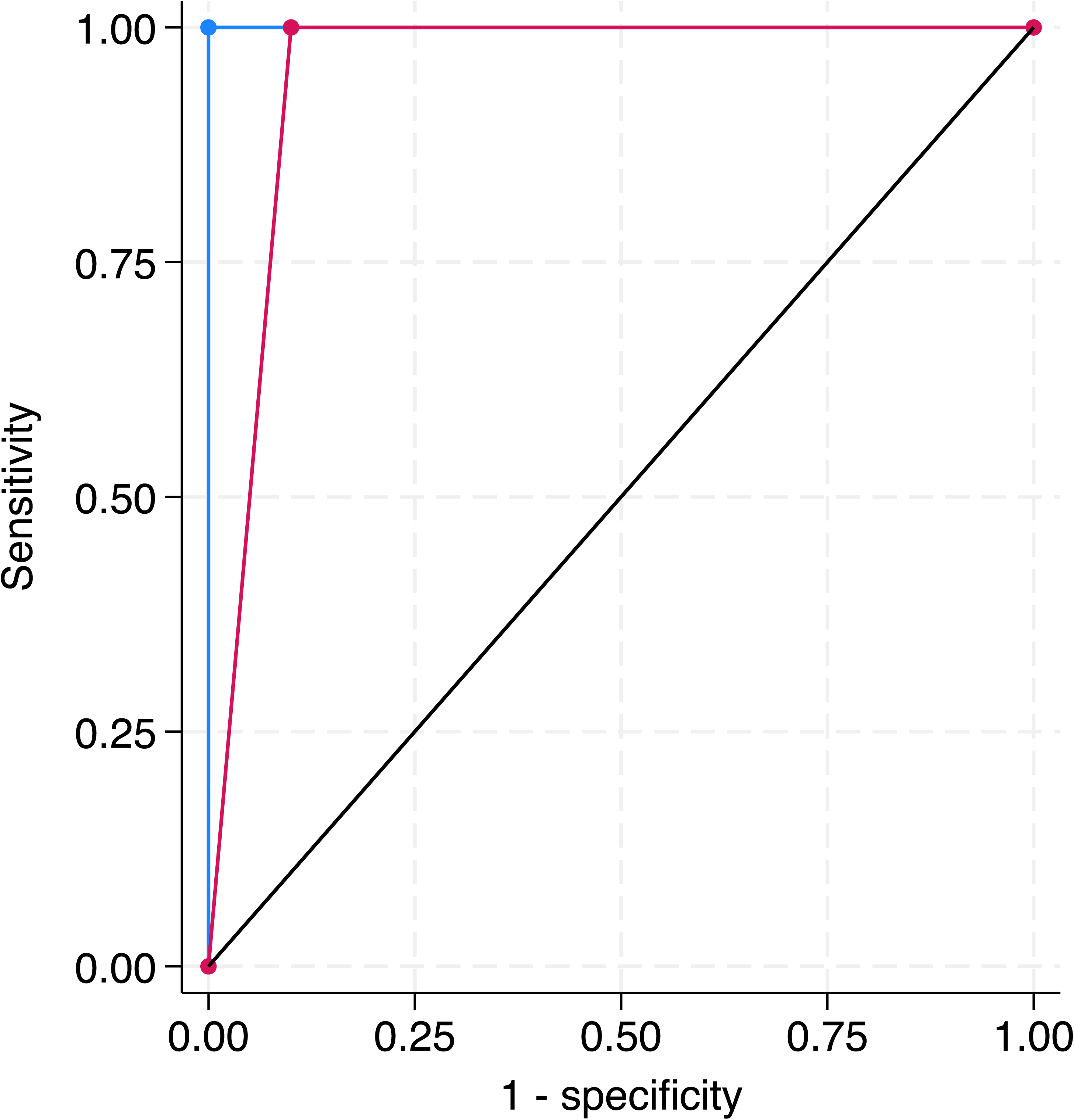
Receiver operating characteristic analysis with area under the curve (AUC) for proposed model and 2018 system: external validation set (10 BWSp cases, 10 controls, 10 overgrowth cases). Proposed model AUC = 1.00 (95% CI, 1.00-1.00); 2018 system AUC = 0.95 (95% CI, 0.88-1.00). Classification performance was not significantly different (χ² = 2.11, *P* = .15).

## Discussion

### Clinical implications

We utilized machine learning to develop a simplified diagnostic tool for BWSp. Development, testing, and external validation demonstrate that a reduced set of features retains substantial diagnostic power. Further, these results reaffirm that macroglossia and lateralized overgrowth are truly cardinal features. The observation that macroglossia is more predictive than lateralized overgrowth is unsurprising, given that BWSp is the leading cause of macroglossia whereas lateralized overgrowth occurs in other overgrowth syndromes and as an isolated finding.^12–14^

Although the 2018 guidelines include six cardinal features and eight suggestive features, our analysis indicates that a six-feature model performs comparably (Figure 3).^2^ The predictive power of this parsimonious approach prompts reassessment of the incremental diagnostic value added by routinely evaluating every feature in the 2018 guidelines. Absence of strong predictors like macroglossia and/or lateralized overgrowth should not preclude further evaluation for BWSp; however, the presence of either or both features should strongly increase clinical suspicion. Careful assessment for subtle macroglossia and lateralized overgrowth is advised in patients presenting with any cardinal or suggestive feature.

Clinicians report that time constraints and complex guidelines create barriers to implementing clinical recommendations in routine medicine—especially when recommendations require the integration of multiple factors.^15^ Our simplified model may be especially useful for physicians with high patient volumes by allowing them to prioritize key findings. Functional diagnostic criteria are also valuable in settings where sensitive molecular testing is unavailable.

Features excluded from our model continue to offer diagnostic clues. For example, ear creases/pits and facial nevus simplex were highly prevalent in our BWSp cohort (Table 1). Although both findings are common in BWSp, they are also frequently appreciated in children without a genetic diagnosis and in those with genetic conditions other than BWSp.^9,16,17^ Nonetheless, their high prevalences in this cohort suggest that examination for strongly predictive features (e.g., macroglossia, lateralized overgrowth) is warranted in children with suggestive features.

Cardinal features beyond lateralized overgrowth, macroglossia, and omphalocele still had diagnostic utility. A review of the model’s missed cases in the testing set showed that several patients, specifically those with pUPD11 and GWpUPD, presented with hyperinsulinism. Hyperinsulinism is highly characteristic of these subtypes and remains an important component of their clinical picture (Supplemental Table S5).^18^ There were three patients whose testing indication was presentation with Wilms tumor; in the absence of a tumor, none would have met 2018 criteria to pursue genetic testing. This underscores the importance of evaluating any child with Wilms tumor, unilateral or multifocal, for BWSp even when other classic features are absent. Notably, both patients with IC1 GOM presented with bilateral Wilms tumors and no additional BWSp features (Supplemental Table S5). This, together with their lower likelihood of clinical diagnosis, suggests that children with IC1 GOM may be at risk of being missed by both the 2018 guidelines and the proposed model, consistent with prior reports that this subgroup often presents with a milder external phenotype.^9–11^ This concern is amplified by the fact that IC1 GOM carries the highest tumor risk.^3^ These missed cases emphasize that the simplified model does not replace clinical judgment or discount other testing indications (e.g., tumors, hyperinsulinism).

Interestingly, developmental delay was a negative predictor of BWSp. This outcome does not mean that developmental concerns are incompatible with workup for BWSp, as this finding could be artifactual and may reflect a referral bias (e.g., patients with developmental delay may be more likely to be seen in genetics clinics). Additional research is needed to clarify whether there is truly an inverse relationship between developmental delay and BWSp.

### Future directions and limitations

This BWSp cohort consists of patients evaluated at a single center, so generalizability may be limited. We partially addressed this concern by validating our model in an external cohort. While the model perfectly classified this set, we do not assume this level of performance will occur universally, especially since differences were observed between development and external data sets (Supplemental Tables S7 and S8). In addition, the external cohort is limited to more common molecular subtypes (IC2 LOM and pUPD11), so model validation in cohorts containing rarer genotypes is necessary. We are actively seeking collaborators willing to contribute additional patient data.

Although the development set included a few controls with overgrowth syndromes, most had no known genetic diagnosis, limiting our ability to evaluate model performance for phenotypically similar conditions. For example, macroglossia is common in Simpson-Golabi-Behmel syndrome (OMIM 312870), and lateralized overgrowth occurs in other disorders, notably PIK3CA-related overgrowth spectrum (MONDO 1040002).^12,19^ This phenotypic overlap could reduce the model’s specificity, so clinical judgment and molecular testing remain important companions to this simplified algorithm. Additional features may be needed to distinguish BWSp from other overgrowth syndromes.

Mosaicism poses several challenges for diagnosis, management, and study of BWSp. Because epigenetic or genetic changes may be confined to only some tissues, or occur at low levels of mosaicism, reliance on blood testing can lead to the under-detection of certain subtypes. In our cohort, patients with IC2 LOM (*P* < .001) and pUPD11 (*P* = .03) were more likely to test positive in blood compared to those with IC1 GOM, and, consistent with prior reports, IC1 GOM cases were more frequently identified through other tissues.^9^ If some subtypes are underreported, the prevalence of milder external phenotypes in these groups may be underestimated.

The large representation of IC2 LOM in our cohort, while mirroring the genotype distribution observed broadly, may bias the model toward features common in this subtype. For example, macroglossia and omphalocele were most prevalent in the IC2 LOM group (Table 1). Moreover, midface flattening was most frequently observed in IC2 LOM (Table 1), raising the possibility that we are describing a facial phenotype skewed towards one subtype. Future work should refine our concept of the BWSp facial gestalt and investigate differences that may exist across subtypes.

While ear creases/pits and macroglossia are the only craniofacial findings included in the 2018 scoring system, this study, which identified midface flattening as a predictor, and advances in image-based machine learning invite the possibility that facial morphology could become a diagnostic tool for BWSp. Recent work with a Swap Disentangled Variational Autoencoder demonstrated that artificial intelligence successfully distinguishes controls from patients with BWSp by identifying characteristic facial morphologies.^20^ Researchers are also exploring whether artificial intelligence can differentiate between genetic syndromes, which could guide targeted testing.^21,22^ As larger patient image datasets are compiled, it will be important to test whether imaging can address gaps in detecting mosaic conditions.

## Conclusion

A simplified six-feature diagnostic model, driven by macroglossia and lateralized overgrowth, identifies molecularly confirmed BWSp cases with performance comparable to the 2018 scoring system. However, clinical judgment and targeted testing for cardinal features or presentation with a tumor remain essential. Independent validation studies should evaluate and refine this model.

## Data Availability

De-identified data may be made available upon request to the corresponding author.

## Acknowledgements

This work is supported by the Lorenzo “Turtle” Sartini Jr. Endowed Chair in Beckwith-Wiedemann Syndrome Research (JMK) and the Cornerstone Fund for Beckwith-Wiedemann Syndrome Research. We would like to thank the patients and their family members for their participation in our study.

## Author Contributions

Conceptualization: J.M.K.; Data curation: S.A.A., A.V., B.T.D., A.M.G., J.A.F.; Formal analysis: S.A.A., D.S., C.M.C., J.M.K.; Funding acquisition: J.M.K.; Investigation: S.A.A., A.V., B.T.D., A.M.G., J.A.F.; Supervision: J.M.K.; Writing-original draft: S.A.A., J.M.K.; Writing-review and editing: A.V., B.T.D., A.M.G., J.A.F., D.S., C.M.C., J.M.K.

## Ethics Declaration

Informed consent, where applicable, was obtained from participants or families according to the relevant IRB protocol. Chart review and data abstraction were performed under IRB13-010658 and IRB19-016459.

## References

1. Mussa A, Russo S, De Crescenzo A, et al. Prevalence of Beckwith-Wiedemann syndrome in North West of Italy. Am J Med Genet A. Oct 2013;161A(10):2481–6. doi:10.1002/ajmg.a.36080

2. Brioude F, Kalish JM, Mussa A, et al. Expert consensus document: Clinical and molecular diagnosis, screening and management of Beckwith-Wiedemann syndrome: an international consensus statement. Nat Rev Endocrinol. Apr 2018;14(4):229–249. doi:10.1038/nrendo.2017.166

3. Kalish JM, Becktell KD, Bougeard G, et al. Update on Surveillance for Wilms Tumor and Hepatoblastoma in Beckwith-Wiedemann Syndrome and Other Predisposition Syndromes. Clin Cancer Res. Dec 2 2024;30(23):5260–5269. doi:10.1158/1078-0432.CCR-24-2100

4. George AM, Viswanathan A, Best LG, et al. Expanded phenotype and cancer risk in patients with Beckwith-Wiedemann spectrum caused by CDKN1C variants. Am J Med Genet A. Oct 2024;194(10):e63777. doi:10.1002/ajmg.a.63777

5. Erwin AL, El Haija AA, Bennett JT, et al. Isolated lateralized overgrowth and the need for tumor screening: A clinical practice resource of the American College of Medical Genetics and Genomics (ACMG). Genet Med. Oct 2025;27(10):101480. doi:10.1016/j.gim.2025.101480

6. Russo S, Calzari L, Mussa A, et al. A multi-method approach to the molecular diagnosis of overt and borderline 11p15.5 defects underlying Silver-Russell and Beckwith-Wiedemann syndromes. Clin Epigenetics. 2016;8:23. doi:10.1186/s13148-016-0183-8

7. Mussa A, Molinatto C, Cerrato F, et al. Assisted Reproductive Techniques and Risk of Beckwith-Wiedemann Syndrome. Pediatrics. Jul 2017;140(1)doi:10.1542/peds.2016-4311

8. Baker EK, Merton CF, Tan WH, Dudding-Byth T, Godler DE, Sadhwani A. Methylation analysis and developmental profile of two individuals with Angelman syndrome due to mosaic imprinting defects. Eur J Med Genet. Apr 2022;65(4):104456. doi:10.1016/j.ejmg.2022.104456

9. Duffy KA, Cielo CM, Cohen JL, et al. Characterization of the Beckwith-Wiedemann spectrum: Diagnosis and management. Am J Med Genet C Semin Med Genet. Dec 2019;181(4):693–708. doi:10.1002/ajmg.c.31740

10. MacFarland SP, Duffy KA, Bhatti TR, et al. Diagnosis of Beckwith-Wiedemann syndrome in children presenting with Wilms tumor. Pediatr Blood Cancer. Oct 2018;65(10):e27296. doi:10.1002/pbc.27296

11. Scott RH, Douglas J, Baskcomb L, et al. Constitutional 11p15 abnormalities, including heritable imprinting center mutations, cause nonsyndromic Wilms tumor. Nat Genet. Nov 2008;40(11):1329–34. doi:10.1038/ng.243

12. Mussa A, Carli D, Cardaropoli S, Ferrero GB, Resta N. Lateralized and Segmental Overgrowth in Children. Cancers (Basel). Dec 7 2021;13(24). doi:10.3390/cancers13246166

13. Prada CE, Zarate YA, Hopkin RJ. Genetic causes of macroglossia: diagnostic approach. Pediatrics. Feb 2012;129(2):e431–7. doi:10.1542/peds.2011-1732

14. Kutti Sridharan G, Rokkam VR. Macroglossia. StatPearls. 2023.

15. Qumseya B, Goddard A, Qumseya A, Estores D, Draganov PV, Forsmark C. Barriers to Clinical Practice Guideline Implementation Among Physicians: A Physician Survey. Int J Gen Med. 2021;14:7591–7598. doi:10.2147/IJGM.S333501

16. Bartel-Friedrich S, Wulke C. Classification and diagnosis of ear malformations. GMS Curr Top Otorhinolaryngol Head Neck Surg. 2007;6:Doc05.

17. Juern AM, Glick ZR, Drolet BA, Frieden IJ. Nevus simplex: a reconsideration of nomenclature, sites of involvement, and disease associations. J Am Acad Dermatol. Nov 2010;63(5):805–14. doi:10.1016/j.jaad.2009.08.066

18. George AM, Viswanathan A, Sussman JH, et al. Determinants of Hyperinsulinism Severity in Children with Beckwith-Wiedemann Syndrome. J Clin Endocrinol Metab. Feb 7 2026. doi:10.1210/clinem/dgag053

19. Nisbet AF, Viswanathan A, George AM, et al. Phenotypic spectrum and tumor risk in Simpson-Golabi-Behmel syndrome: Case series and comprehensive literature review. Am J Med Genet A. Dec 2024;194(12):e63840. doi:10.1002/ajmg.a.63840

20. Rijlaarsdam T, Smith L, Rickart A, et al. The Identification of Beckwith-Wiedemann Syndrome Through Swap Disentangled Variational Autoencoder. J Craniofac Surg. Mar 10 2026. doi:10.1097/SCS.0000000000012540

21. Hallgrimsson B, Aponte JD, Katz DC, et al. Automated syndrome diagnosis by three-dimensional facial imaging. Genet Med. Oct 2020;22(10):1682–1693. doi:10.1038/s41436-020-0845-y

22. Wu D, Yang J, Liu C, et al. GestaltMML: Enhancing Rare Genetic Disease Diagnosis through Multimodal Machine Learning Combining Facial Images and Clinical Texts. Preprint. *ArXiv*. Apr 22 2024;arXiv:2312.15320v2.

