## Supplemental Table S1 for "Machine-Learning Model Identifies New Diagnostic Criteria for Beckwith-Wiedemann Spectrum"

Supplemental Table S1. Demographics: BWSp Cohort

| **Characteristic** | **Total, n/N (%)** |
| --- | --- |
| Sex, male | 234/549 (42.6) |
| Lives in United States | 488/555 (87.9) |
| Molecular subtype |  |
| IC1 GOM | 67/555 (12.1) |
| IC2 LOM | 295/555 (53.2) |
| pUPD11 | 141/555 (25.4) |
| *CDKN1C* variant | 14/555 (2.5) |
| GWpUPD | 9/555 (1.6) |
| 11p15 anomaly | 27/555 (4.9) |
| Other^a^ | 2/555 (0.4) |

^a^ “Other” category includes one patient with IC2 LOM/IC1 LOM (BWSp/Russell-Silver syndrome) and one patient with IC1 GOM/CDKN1C.
