## Supplemental Table S2 for "Machine-Learning Model Identifies New Diagnostic Criteria for Beckwith-Wiedemann Spectrum"

Supplemental Table S2. Clinical Features of the Control Cohort^a^

|  | **Total** | **No genetic condition** | **Non-overgrowth genetic condition**^b^ | **Non-BWSp genetic overgrowth condition**^c^ |
| --- | --- | --- | --- | --- |
| **Cardinal Features** |  |  |  |  |
| Macroglossia | 12/150 (8) | 11/128 (9) | 1/13 (8) | 0/9 (0) |
| Omphalocele | 4/150 (3) | 4/128 (3) | 0/13 (0) | 0/9 (0) |
| Lateralized overgrowth | 19/150 (13) | 15/128 (12) | 2/13 (15) | 2/9 (22) |
| Multifocal/bilateral WT/NB | 2/150 (1) | 2/128 (2) | 0/13 (0) | 0/9 (0) |
| Hyperinsulinism | 10/150 (7) | 4/128 (3) | 6/13 (46) | 0/9 (0) |
| Pathology findings^d^ | 1/143 (1) | 1/121 (1) | 0/13 (0) | 0/9 (0) |
| **Suggestive Features** |  |  |  |  |
| Large for gestational age | 23/149 (15) | 14/127 (11) | 5/13 (38) | 4/9 (44) |
| Facial nevus simplex | 27/150 (18) | 23/128 (18) | 3/13 (23) | 1/9 (11) |
| Polyhydramnios | 10/150 (7) | 8/128 (6) | 1/13 (8) | 1/9 (11) |
| Placentomegaly | 1/150 (1) | 1/128 (1) | 0/13 (0) | 0/9 (0) |
| Ear creases/pits | 31/150 (21) | 26/128 (20) | 3/13 (23) | 2/9 (22) |
| Transient hypoglycemia | 20/150 (13) | 17/128 (13) | 2/13 (15) | 1/9 (11) |
| Tumor^e^ | 17/150 (11) | 15/128 (12) | 1/13 (8) | 1/9 (11) |
| Wilms tumor^f^ | 11/150 (7) | 11/128 (9) | 0/13 (0) | 0/9 (0) |
| Hepatoblastoma | 2/150 (1) | 2/128 (2) | 0/13 (0) | 0/9 (0) |
| Organomegaly |  |  |  |  |
| Nephromegaly | 5/150 (3) | 5/128 (4) | 0/13 (0) | 0/9 (0) |
| Hepatomegaly | 8/150 (5) | 6/128 (5) | 1/13 (8) | 1/9 (11) |
| Abdominal wall defect |  |  |  |  |
| Umbilical hernia | 29/150 (19) | 21/128 (16) | 3/13 (23) | 5/9 (56) |
| Diastasis recti | 13/150 (9) | 12/128 (9) | 0/13 (0) | 1/9 (11) |
| **Other Features** |  |  |  |  |
| Assisted reproductive technology | 15/142 (11) | 14/121 (12) | 0/12 (0) | 1/9 (11) |
| Midface flattening | 6/150 (4) | 6/128 (5) | 0/13 (0) | 0/9 (0) |
| Thin upper lip | 16/150 (11) | 13/128 (10) | 3/13 (23) | 0/9 (0) |
| Cardiac anomalies | 20/150 (13) | 13/128 (10) | 6/13 (46) | 1/9 (11) |
| Developmental delay | 73/150 (49) | 58/128 (45) | 11/13 (85) | 4/9 (44) |
| Prematurity (< 37 weeks) | 26/146 (18) | 23/124 (19) | 2/13 (15) | 1/9 (11) |
| Gestational age, mean (SD), weeks | 38.3 (2.1) | 38.3 (2.1) | 37.8 (0.3) | 38.1 (0.2) |

Abbreviations: WT/NB, Wilms tumor or nephroblastomatosis.

^a^ Categorical variables reported as n/N (%), where N excludes patients with unknown or unavailable data.

^b^ Non-overgrowth genetic conditions include 22q11.2 deletion syndrome (1), *DNAH11* variant (1), F2 mutation (1), G6PD deficiency (1), Gaucher disease (1), *GDF5* variant (1), Kabuki syndrome (2), monogenic hyperinsulinism (4), ReNU syndrome (1).

^c^ Non-BWSp overgrowth conditions include 48 XXYY syndrome (1), Costello syndrome (1), neurofibromatosis type 1 (1), PIK3CA-related overgrowth spectrum (3), PTEN hamartoma tumor syndrome (1), SUZ12-related overgrowth disorder (1), *TSC2* variant (1).

^d^ Placental mesenchymal dysplasia, adenomatosis of the pancreas, or cytomegaly of the adrenal glands.

^e^ Tumors observed in this cohort include hepatoblastoma, neuroblastoma, rhabdomyosarcoma, teratoma, multifocal/bilateral Wilms tumor, and unilateral Wilms tumor.

^f^ Wilms tumor category includes both multifocal/bilateral and unilateral cases.
