## Supplemental Table S3 for "Machine-Learning Model Identifies New Diagnostic Criteria for Beckwith-Wiedemann Spectrum"

Supplemental Table S3. Comparison of Development and Testing Sets: BWSp Cases^a^

|  | **Development** | **Testing** | ***P* value** |
| --- | --- | --- | --- |
| **Cardinal Features** |  |  |  |
| Macroglossia | 300/389 (77.1) | 128/166 (77) | > .99 |
| Omphalocele | 64/389 (16.5) | 28/166 (17) | .90 |
| Lateralized overgrowth | 275/389 (70.7) | 108/166 (65) | .19 |
| Multifocal/bilateral WT/NB | 10/389 (2.6) | 4/166 (2) | > .99 |
| Hyperinsulinism | 62/389 (15.9) | 34/166 (20) | .20 |
| Pathology findings^b^ | 13/355 (3.7) | 4/150 (3) | .57 |
| **Suggestive Features** |  |  |  |
| Large for gestational age | 138/385 (35.8) | 69/166 (42) | .20 |
| Facial nevus simplex | 187/369 (50.7) | 77/159 (48) | .64 |
| Polyhydramnios | 98/380 (25.8) | 39/163 (24) | .65 |
| Placentomegaly | 38/365 (10.4) | 12/157 (8) | .32 |
| Ear creases/pits | 224/389 (57.6) | 101/166 (61) | .48 |
| Transient hypoglycemia | 135/389 (34.7) | 55/165 (33) | .76 |
| Tumor |  |  |  |
| Wilms tumor^c^ | 29/389 (7.5) | 11/166 (7) | .73 |
| Hepatoblastoma | 14/389 (3.6) | 7/166 (4) | .73 |
| Organomegaly |  |  |  |
| Nephromegaly | 60/384 (15.6) | 31/164 (19) | .35 |
| Hepatomegaly | 70/384 (18.2) | 32/164 (20) | .72 |
| Abdominal wall defect |  |  |  |
| Umbilical hernia | 135/389 (34.7) | 68/165 (41) | .15 |
| Diastasis recti | 94/388 (24.2) | 40/165 (24) | > .99 |
| **Other Features** |  |  |  |
| Assisted reproductive technology | 75/314 (23.9) | 29/135 (21) | .58 |
| Midface flattening | 109/354 (30.8) | 50/151 (33) | .61 |
| Thin upper lip | 102/354 (28.8) | 44/151 (29) | .94 |
| Cardiac anomalies | 117/384 (30.5) | 49/165 (30) | .86 |
| Developmental delay | 169/367 (46.0) | 79/157 (50) | .37 |
| Prematurity (< 37 weeks) | 157/371 (42.3) | 62/159 (39) | .48 |
| Gestational age, mean (SD), weeks | 36.3 (3.7) | 36.7 (3.7) | .19 |

Abbreviations: WT/NB, Wilms tumor or nephroblastomatosis.

^a^ Categorical variables reported as n/N (%), where N excludes patients with unknown or unavailable data.

^b^ Placental mesenchymal dysplasia, adenomatosis of the pancreas, or cytomegaly of the adrenal glands.

^c^ Wilms tumor category includes both multifocal/bilateral and unilateral cases.
