## Supplemental Table S4 for "Machine-Learning Model Identifies New Diagnostic Criteria for Beckwith-Wiedemann Spectrum"

Supplemental Table S4. Comparison of Development and Testing Sets: Controls^a^

|  | **Development** | **Testing** | ***P* value** |
| --- | --- | --- | --- |
| **Cardinal Features** |  |  |  |
| Macroglossia | 9/105 (9) | 3/45 (7) | > .99 |
| Omphalocele | 4/105 (4) | 0/45 (0) | .32 |
| Lateralized overgrowth | 15/105 (14) | 4/45 (9) | .36 |
| Multifocal/bilateral WT/NB | 2/105 (2) | 0/45 (0) | > .99 |
| Hyperinsulinism | 6/105 (6) | 4/45 (9) | .49 |
| Pathology findings^b^ | 1/101 (1) | 0/42 (0) | > .99 |
| **Suggestive Features** |  |  |  |
| Large for gestational age | 20/104 (19) | 3/45 (7) | .05 |
| Facial nevus simplex | 15/105 (14) | 12/45 (27) | .07 |
| Polyhydramnios | 6/105 (6) | 4/45 (9) | .49 |
| Placentomegaly | 0/105 (0) | 1/45 (2) | .30 |
| Ear creases/pits | 16/105 (15) | 15/45 (33) | .01 |
| Transient hypoglycemia | 16/105 (15) | 4/45 (9) | .29 |
| Tumor |  |  |  |
| Wilms tumor^c^ | 8/105 (8) | 3/45 (7) | > .99 |
| Hepatoblastoma | 1/105 (1) | 1/45 (2) | .51 |
| Organomegaly |  |  |  |
| Nephromegaly | 3/105 (3) | 2/45 (4) | .64 |
| Hepatomegaly | 7/105 (7) | 1/45 (2) | .44 |
| Abdominal wall defect |  |  |  |
| Umbilical hernia | 23/105 (22) | 6/45 (13) | .22 |
| Diastasis recti | 5/105 (5) | 8/45 (18) | .02 |
| **Other Features** |  |  |  |
| Assisted reproductive technology | 10/99 (10) | 5/43 (12) | .77 |
| Midface flattening | 4/105 (4) | 2/45 (4) | > .99 |
| Thin upper lip | 10/105 (10) | 6/45 (13) | .57 |
| Cardiac anomalies | 17/105 (16) | 3/45 (7) | .12 |
| Developmental delay | 56/105 (53) | 17/45 (38) | .08 |
| Prematurity (< 37 weeks) | 22/102 (22) | 4/44 (9) | .07 |
| Gestational age, mean (SD), weeks | 38.1 (2.1) | 38.8 (2.1) | .02 |

Abbreviations: WT/NB, Wilms tumor or nephroblastomatosis.

^a^ Categorical variables reported as n/N (%), where N excludes patients with unknown or unavailable data.

^b^ Placental mesenchymal dysplasia, adenomatosis of the pancreas, or cytomegaly of the adrenal glands.

^c^ Wilms tumor category includes both multifocal/bilateral and unilateral cases.
