## Supplemental Table S5 for "Machine-Learning Model Identifies New Diagnostic Criteria for Beckwith-Wiedemann Spectrum"

Supplemental Table S5. Review of BWSp Cases Missed by the Proposed Model

| **Subject ID** | **Indication for genetic testing** | **Clinical score** | **Clinical score excluding tumor** | **Tumor** | **Cardinal features** | **Suggestive features** |
| --- | --- | --- | --- | --- | --- | --- |
| **IC1 GOM** |  |  |  |  |  |  |
| 1 | Presented with tumor | 2 | 0 | Bilateral Wilms | — | — |
| 2 | Presented with tumor | 2 | 0 | Bilateral Wilms | — | — |
| **IC2 LOM** |  |  |  |  |  |  |
| 3 | Suggestive features and subtle thigh asymmetry | 2 | 2 | — | — | Facial nevus simplex, ear creases |
| 4 | Suspicion for Russell-Silver syndrome | 4 | 4 | — | Lateralized overgrowth | Transient hypoglycemia, diastasis recti |
| 5 | Cardinal feature | 3 | 3 | — | Macroglossia | Umbilical hernia |
| 6 | Cardinal feature | 5 | 5 | — | Macroglossia | Polyhydramnios, ear creases, umbilical hernia |
| 7 | Familial concern | 2 | 2 | — | — | LGA, facial nevus simplex |
| 8 | Cardinal feature | 4 | 4 | — | Omphalocele | LGA, ear crease |
| 9 | Presented with tumor | 2 | 1 | Unilateral Wilms | — | Umbilical hernia |
| 10^a^ | Unknown | 2 | 2 | — | — | LGA, transient hypoglycemia |
| 11 | Cardinal feature | 3 | 3 | — | Lateralized overgrowth | LGA |
| **pUPD11** |  |  |  |  |  |  |
| 12 | Cardinal feature | 7 | 7 | — | Lateralized overgrowth | LGA, placentomegaly, ear creases, transient hypoglycemia, umbilical hernia, diastasis recti |
| 13 | Cardinal feature | 6 | 6 | — | Hyperinsulinism, lateralized overgrowth | LGA, umbilical hernia |
| 14 | Cardinal feature | 2 | 2 | — | Lateralized overgrowth | — |
| 15 | Cardinal feature | 4 | 3 | Hepatoblastoma | Hyperinsulinism | Nephromegaly, hepatomegaly |
| **GWpUPD** |  |  |  |  |  |  |
| 16 | Suspicion of tumor and cardinal feature | 6 | 6 | — | Hyperinsulinism | Polyhydramnios, ear creases, hepatomegaly, umbilical hernia |
| **11p15 anomaly** |  |  |  |  |  |  |
| 17 | Respiratory distress at birth | 5 | 3 | Nephroblastomatosis | Macroglossia | Umbilical hernia |

^a^ External subject reported in literature.
