## Supplemental Table S6 for "Machine-Learning Model Identifies New Diagnostic Criteria for Beckwith-Wiedemann Spectrum"

Supplemental Table S6. Clinical Features of the External Cohort^a^

|  | **BWSp** | **Controls** | **Non-BWSp genetic overgrowth condition** |
| --- | --- | --- | --- |
| **Cardinal Features** |  |  |  |
| Macroglossia | 7/10 (70) | 0/10 (0) | 1/10 (10) |
| Omphalocele | 5/10 (50) | 0/10 (0) | 0/10 (0) |
| Lateralized overgrowth | 4/10 (40) | 0/10 (0) | 1/10 (10) |
| Multifocal/bilateral WT/NB | 0/10 (0) | 0/10 (0) | 0/10 (0) |
| Hyperinsulinism | 0/8 (0) | 0/7 (0) | 0/9 (0) |
| Pathology findings^b^ | 2/5 (40) | 0/0 | 0/0 |
| **Suggestive Features** |  |  |  |
| Large for gestational age | 6/9 (67) | 0/9 (0) | 2/10 (20) |
| Facial nevus simplex | 6/9 (67) | 1/10 (10) | 1/9 (11) |
| Polyhydramnios | 7/8 (88) | 0/9 (0) | 1/9 (11) |
| Placentomegaly | 2/4 (50) | 0/0 | 0/0 |
| Ear creases/pits | 9/10 (90) | 1/10 (10) | 2/9 (22) |
| Transient hypoglycemia | 6/10 (60) | 5/10 (50) | 1/9 (11) |
| Tumor |  |  |  |
| Wilms tumor^c^ | 1/10 (10) | 0/10 (0) | 0/10 (0) |
| Hepatoblastoma | 0/10 (0) | 0/10 (0) | 0/10 (0) |
| Organomegaly |  |  |  |
| Nephromegaly | 3/10 (30) | 0/8 (0) | 2/7 (29) |
| Hepatomegaly | 3/10 (30) | 0/9 (0) | 0/5 (0) |
| Abdominal wall defect |  |  |  |
| Umbilical hernia | 2/9 (22) | 0/10 (0) | 1/9 (11) |
| Diastasis recti | 1/9 (11) | 0/10 (0) | 1/10 (10) |
| **Other Features** |  |  |  |
| Assisted reproductive technology | 1/5 (20) | 1/10 (10) | 1/5 (20) |
| Midface flattening | 8/8 (100) | 5/10 (50) | 1/10 (10) |
| Thin upper lip | 1/1 (100) | 4/9 (44) | 2/10 (20) |
| Cardiac anomalies | 8/10 (80) | 3/10 (30) | 5/9 (56) |
| Developmental delay | 2/10 (20) | 10/10 (100) | 10/10 (100) |

Abbreviations: WT/NB, Wilms tumor or nephroblastomatosis.

^a^ Categorical variables reported as n/N (%), where N excludes patients with unknown or unavailable data.

^b^ Placental mesenchymal dysplasia, adenomatosis of the pancreas, or cytomegaly of the adrenal glands.

^c^ Wilms tumor category includes both multifocal/bilateral and unilateral cases.
