## Supplemental Table S7 for "Machine-Learning Model Identifies New Diagnostic Criteria for Beckwith-Wiedemann Spectrum"

Supplemental Table S7. Comparison of Development and External Sets: BWSp Cases^a^

|  | **Development** | **External** | ***P* value** |
| --- | --- | --- | --- |
| **Cardinal Features** |  |  |  |
| Macroglossia | 300/389 (77.1) | 7/10 (70) | .70 |
| Omphalocele | 64/389 (16.5) | 5/10 (50) | .02 |
| Lateralized overgrowth | 275/389 (70.7) | 4/10 (40) | .07 |
| Multifocal/bilateral WT/NB | 10/389 (2.6) | 0/10 (0) | > .99 |
| Hyperinsulinism | 62/389 (15.9) | 0/8 (0) | .62 |
| Pathology findings^b^ | 13/355 (3.7) | 2/5 (40) | .02 |
| **Suggestive Features** |  |  |  |
| Large for gestational age | 138/385 (35.8) | 6/9 (67) | .08 |
| Facial nevus simplex | 187/369 (50.7) | 6/9 (67) | .50 |
| Polyhydramnios | 98/380 (25.8) | 7/8 (88) | < .001 |
| Placentomegaly | 38/365 (10.4) | 2/4 (50) | .06 |
| Ear creases/pits | 224/389 (57.6) | 9/10 (90) | .05 |
| Transient hypoglycemia | 135/389 (34.7) | 6/10 (60) | .18 |
| Tumor |  |  |  |
| Wilms tumor^c^ | 29/389 (7.5) | 1/10 (10) | .55 |
| Hepatoblastoma | 14/389 (3.6) | 0/10 (0) | > .99 |
| Organomegaly |  |  |  |
| Nephromegaly | 60/384 (15.6) | 3/10 (30) | .20 |
| Hepatomegaly | 70/384 (18.2) | 3/10 (30) | .40 |
| Abdominal wall defect |  |  |  |
| Umbilical hernia | 135/389 (34.7) | 2/9 (22) | .72 |
| Diastasis recti | 94/388 (24.2) | 1/9 (11) | .69 |
| **Other Features** |  |  |  |
| Assisted reproductive technology | 75/314 (23.9) | 1/5 (20) | > .99 |
| Midface flattening | 109/354 (30.8) | 8/8 (100) | < .001 |
| Thin upper lip | 102/354 (28.8) | 1/1 (100) | .29 |
| Cardiac anomalies | 117/384 (30.5) | 8/10 (80) | .002 |
| Developmental delay | 169/367 (46.0) | 2/10 (20) | .12 |

Abbreviations: WT/NB, Wilms tumor or nephroblastomatosis.

^a^ Categorical variables reported as n/N (%), where N excludes patients with unknown or unavailable data.

^b^ Placental mesenchymal dysplasia, adenomatosis of the pancreas, or cytomegaly of the adrenal glands.

^c^ Wilms tumor category includes both multifocal/bilateral and unilateral cases.
