## Supplemental Table S8 for "Machine-Learning Model Identifies New Diagnostic Criteria for Beckwith-Wiedemann Spectrum"

Supplemental Table S8. Comparison of Development and External Sets: Controls^a^

|  | **Development** | **External** | ***P* value** |
| --- | --- | --- | --- |
| **Cardinal Features** |  |  |  |
| Macroglossia | 9/105 (9) | 0/10 (0) | > .99 |
| Omphalocele | 4/105 (4) | 0/10 (0) | > .99 |
| Lateralized overgrowth | 15/105 (14) | 0/10 (0) | .36 |
| Multifocal/bilateral WT/NB | 2/105 (2) | 0/10 (0) | > .99 |
| Hyperinsulinism | 6/105 (6) | 0/7 (0) | > .99 |
| Pathology findings^b^ | 1/101 (1) | 0/0 | — |
| **Suggestive Features** |  |  |  |
| Large for gestational age | 20/104 (19) | 0/9 (0) | .36 |
| Facial nevus simplex | 15/105 (14) | 1/10 (10) | > .99 |
| Polyhydramnios | 6/105 (6) | 0/9 (0) | > .99 |
| Placentomegaly | 0/105 (0) | 0/0 | — |
| Ear creases/pits | 16/105 (15) | 1/10 (10) | > .99 |
| Transient hypoglycemia | 16/105 (15) | 5/10 (50) | .02 |
| Tumor |  |  |  |
| Wilms tumor^c^ | 8/105 (8) | 0/10 (0) | > .99 |
| Hepatoblastoma | 1/105 (1) | 0/10 (0) | > .99 |
| Organomegaly |  |  |  |
| Nephromegaly | 3/105 (3) | 0/8 (0) | > .99 |
| Hepatomegaly | 7/105 (7) | 0/9 (0) | > .99 |
| Abdominal wall defect |  |  |  |
| Umbilical hernia | 23/105 (22) | 0/10 (0) | .21 |
| Diastasis recti | 5/105 (5) | 0/10 (0) | > .99 |
| **Other Features** |  |  |  |
| Assisted reproductive technology | 10/99 (10) | 1/10 (10) | > .99 |
| Midface flattening | 4/105 (4) | 5/10 (50) | < .001 |
| Thin upper lip | 10/105 (10) | 4/9 (44) | .01 |
| Cardiac anomalies | 17/105 (16) | 3/10 (30) | .37 |
| Developmental delay | 56/105 (53) | 10/10 (100) | .005 |

Abbreviations: WT/NB, Wilms tumor or nephroblastomatosis.

^a^ Categorical variables reported as n/N (%), where N excludes patients with unknown or unavailable data.

^b^ Placental mesenchymal dysplasia, adenomatosis of the pancreas, or cytomegaly of the adrenal glands.

^c^ Wilms tumor category includes both multifocal/bilateral and unilateral cases.
